# PARP1/2 imaging with ^18^F-PARPi in patients with head and neck cancer

**DOI:** 10.1101/19009381

**Authors:** Heiko Schöder, Paula Demétrio De Souza França, Reiko Nakajima, Eva Burnazi, Sheryl Roberts, Christian Brand, Milan Grkovski, Audrey Mauguen, Mark P. Dunphy, Ronald A. Ghossein, Serge Lyashchenko, Jason S. Lewis, Joseph A. O’Donoghue, Ian Ganly, Snehal G. Patel, Nancy Y. Lee, Thomas Reiner

**Author notes:** Address correspondence to: Heiko Schöder, MD, Department of Radiology, Memorial Sloan Kettering Cancer Center, 1275 York Avenue, New York, NY, 10065., Or to: Thomas Reiner, PhD, Department of Radiology and Chemical Biology Program, Memorial Sloan Kettering Cancer Center, 1275 York Avenue, New York, NY, 10065. **Translational Relevance.** Preclinically, labeled PARP1-targeted olaparib derivatives have been used to visualize several malignancies with high contrast, including head and neck cancer. These results suggest that PARP1-targeted imaging agents could potentially be used as a quantitative whole body imaging test for primary and metastatic lesions, improving diagnostic sensitivity and specificity compared to the standard of care. This first-in-human study of ^18^F-PARP1 in patients with head and neck cancer established that imaging with the olaparib-based PARP1 imaging agent ^18^F-PARPi is feasible and safe, and that contrast ratios in the head and neck region are comparable to ^18^F-FDG. Retention in tumors and metastatic nodes is longer than in physiological tissues, including the salivary glands. The lymph node detection rate for ^18^F-PARPi is higher than for ^18^F-FDG, and a subset of ^18^F-PARPi positive and ^18^F-FDG negative lymph nodes resolved after chemoradiation. Further study of ^18^F-PARPi in head and neck cancer is being pursued. ***Funding.*** This work was supported in part by National Institutes of Health grants R01 CA204441, R35 CA232130 and P30 CA008748, the Tow Foundation, the MSK Center for Molecular Imaging & Nanotechnology, the MSK Imaging and Radiation Sciences Program and the MSK Molecularly Targeted Intraoperative Imaging Fund.

## Abstract

**Purpose:** We performed a first-in-human clinical trial. The aim of this study was to determine safety and feasibility of PET imaging with ^18^F-PARPi in patients with head and neck cancer.

**Patients and Methods:** Eleven patients (age 49 to 86 years) with newly diagnosed or recurrent oral and oropharyngeal cancer were injected intravenously with ^18^F-PARPi (331 ± 42 MBq) and dynamic PET/CT imaging was performed between 0 min and 25 min post-injection. Static PET/CT scans were obtained at 30 min, 60 min and 120 min p.i. Blood samples for tracer concentration and metabolite analysis were collected. Blood pressure, ECG, oxygen levels, clinical chemistry and CBC were obtained before and after administration of ^18^F-PARPi.

**Results:** ^18^F-PARPi was well-tolerated by all patients without any safety concerns. Of the 11 patients included in the analysis, ^18^F-PARPi had focal uptake in all primary lesions (n = 10, SUV_max_ = 2.8 ± 1.2) and all ^18^F-FDG positive lymph nodes (n = 34). ^18^F-PARPi uptake was seen in ^18^F-FDG negative lymph nodes of three patients (n = 6). Focal uptake of tracer in primary and metastatic lesions was corroborated by CT alone or in combination with ^18^F-FDG. Contrast for ^18^F-PARPi and ^18^F-FDG was comparable (SUV_max_(lesion)/SUV_max_(genioglossus) = 3.3 and 3.0, respectively; p = 0.23), and SUV_max_ values for ^18^F-PARPi were less variable compared to ^18^F-FDG (1.3 versus 6.0, p = 0.001).

**Conclusions:** Imaging of head and neck cancer with ^18^F-PARPi is feasible and safe. ^18^F-PARPi detects primary and metastatic lesions, and retention in tumors is longer than in healthy tissues.

## Introduction

Diagnosis and treatment of many malignant tumors have dramatically improved in recent decades, and oral and oropharyngeal squamous cell carcinoma are no exception to this trend. However, most patients with oral and oropharyngeal squamous cell carcinoma still present with advanced disease and regional or distant metastases at the time of diagnosis.

Presence of pathological regional lymph nodes is the most powerful and consistent predictor of outcome for oral cancers (1), and the ability to accurately assign the exact extent of metastatic spread within the neck lymphatic system is therefore of great significance (1,2). Complete surgical removal of metastatic disease in the neck has a clear impact on prognosis, but it often remains unclear until histopathological analysis is complete, if a lymph node was metastatic or not (3,4). Clinical palpation of the neck is inadequate, as are the available non-invasive radiological investigative tools. Many patients present with enlarged lymph nodes due to inflammation around the tumor site. These inflamed lymph nodes may mimic neck metastases on CT scans and are often ^18^F-FDG PET avid (5–8). On the other hand, some metastatic neck nodes may not be enlarged and show no abnormal ^18^F-FDG uptake. For these reasons, elective neck dissection or irradiation are often recommended in patients with head and neck cancer for prophylactic treatment of occult metastases. Ideally, however, these procedures, which can lead to increased co-morbidities and reduced quality of life due to overtreatment (9–11), would be avoided if the presence of occult metastases could be definitively ruled out.

PARP1/2 PET imaging could provide a non-invasive solution for this unmet clinical need. PARP1/2 is overexpressed in many malignancies, including oral and oropharyngeal squamous cell carcinoma (12,13), likely due to the enzyme’s central role in DNA repair (14,15). Several radiolabeled PARP1/2 inhibitors were tested in preclinical studies (16,17), showing correlation of uptake with PARP1 expression (18,19), and suggesting that imaging is possible with little unspecific uptake in healthy head and neck tissue.

The purpose of this study was to clinically translate ^18^F-PARPi, a PARP1/2 inhibitor derived from the core scaffold of olaparib (20), and to provide a first step toward validating the tracer as a clinical tool. Other PARP inhibitor-based imaging agents, based on different core scaffolds, were translated earlier (21–23). In this first-in-human phase I clinical trial in patients with oral and oropharyngeal cancer, we determined the safety and pharmacokinetics of ^18^F-PARPi, and correlated its uptake in tumors and normal tissue to standard of care ^18^F-FDG imaging.

## Patients and Methods

### Study design

This exploratory, phase I, single-center, open-label, prospective Health Insurance Portability and Accountability Act (HIPAA) compliant study was approved by the Memorial Sloan Kettering (MSK) Institutional Review Board and written informed consent was obtained from all patients (Clinicaltrials.gov NCT03631017). The primary objectives of this phase I trial were to evaluate the safety of ^18^F-PARPi, to describe the biodistribution and radiation dosimetry, and to describe the tumor uptake of ^18^F-PARPi. Patients were accrued between January 2019 and September 2019 and referred to the MSK Molecular Imaging and Therapy Service for their newly diagnosed or recurrent oral or oropharyngeal cancer. In total, 8 patients with oropharyngeal squamous cell carcinoma and 3 patients with oral cavity squamous cell carcinoma completed the protocol. All 11 patients were injected with ^18^F-PARPi and underwent imaging. Inclusion and exclusion criteria are summarized in Supplementary Table S1.

### Radiopharmaceutical Preparation

^18^F-PARPi was produced under good manufacturing practice (GMP) conditions at the MSK Radiopharmacy under IND# 139,974 similar to previously reported procedures (18-20,24). Briefly, to obtain ^18^F-PARPi, cyclotron-produced [^18^F] was trapped on the QMA SEP pack, eluted with 4 mg of tetraethylammonium bicarbonate into a reaction vial containing a teflon magnetic stir bar, and dried by being heated at 90 °C under vacuum. Then, 2 mg of ethyl-4 nitrobenzoate, dissolved in 0.2 mL DMSO, was added to the reaction vessel, and the reaction was heated to 150 °C for 15 minutes and allowed to cool before 50 μL of 1 M NaOH was added, followed by 50 μL of 1 M HCl, yielding 4-[^18^F]fluorobenzoic acid. Thereafter, 4 mg of 4-(4-fluoro-3-(piperazine-1-carbonyl)benzyl) phthalazin-1(2H)-one in 100 μL dry of DMSO were added followed by 10 mg of HBTU dissolved in 100 μL of DMSO and 20 μL of Et_3_N. ^18^F-PARPi was isolated by preparatory high performance liquid chromatography (HPLC), with isocratic 35% acetonitrile in 0.1% TFA solution as mobile phase, Phenomenex; 6-Phenyl, 5 µm, 250 × 10 mm column as stationary phase at a flow rate of 4.0 mL/min, and a UV detector wavelength of 254 nm. The HPLC system used was a Shimadzu Prominence 20 Series, equipped with a UV detector and a Flow-Ram sodium iodide radioactivity detector supplied by Lablogic, UK. The radiochemical purity range in the ^18^F-PARPi manufactured batches was > 99% (n = 9) and the molar activity was 115 ± 48 GBq/μmol (3.1 ± 1.3 Ci/μmol, n = 9).

### Tracer formulation and quality control

After radiochemical synthesis and purification, the HPLC fraction containing the drug substance was collected at 35 ± 3 min into a pear-shaped glass flask, containing 40 mL of sterile water for injection, USP. To remove the HPLC mobile phase solvents, the reaction mixture was loaded onto a light C18 cartridge and washed with an additional 10 mL of sterile water. The drug substance was then eluted from the C18 SEP Pak with 0.5 mL of ethanol, followed by 9.5 mL of normal saline, through a 0.22 µm sterilizing filter into a sterile, apyrogenic, USP Type I glass vial, sealed with a rubber septum and crimped with an aluminum crimp. The final drug product underwent quality control testing prior to batch release for patient administration in accordance with acceptance specifications for radiochemical purity, endotoxin content, sterilizing filter integrity, pH, appearance, and radionuclide identity confirmation. Sterility testing was performed post-release.

### Procedures

All patients underwent clinical examination, baseline vital signs, pulse oximetry, ECG and blood tests (< 2 weeks prior to imaging). No fasting was required prior to ^18^F-PARPi imaging. On the day of imaging, two IV catheters were placed in each forearm, one for injection of ^18^F-PARPi and one for drawing of IV blood samples. ^18^F-PARPi was administered to patients at an average activity of 333 ± 44 MBq (9.0 ± 1.2 mCi) by intravenous bolus injection. The synthesis of ^18^F-PARPi is summarized in Supplementary Figure S1. For the first six patients, a dynamic PET scan (with the field of view including the heart, lungs, liver, and kidneys) was acquired to study the biodistribution and clearance of the tracer. For the subsequent five patients, the dynamic PET scan was centered on the head and neck region. Immediately after the dynamic study, a static PET/CT scan of the body (extending from skull vertex to upper thighs) was obtained (30 mins p.i.). Two further static PET/CT scans were taken at approximately 60 min and 120 min post-injection. A total of 5 blood samples (at approximately 1 min, 5 min, 30 min, 90 min and 150 min p.i.) were drawn to quantify blood pool activity and to study ^18^F-PARPi metabolites. After imaging, vital signs were obtained and an ECG performed, and blood samples were collected for hematology and blood chemistry analysis). A follow-up phone interview (1–3 days after the imaging study) was conducted to document any side effects occurring after completion of the imaging study.

### Metabolite analysis

Blood clearance measurements were performed as previously reported (25). Briefly, multiple venous blood samples were obtained between 1 min and 150 min after intravenous injection of ^18^F-PARPi. Activity in whole blood and plasma was measured in duplicate using a calibrated NaI (Tl) Wallac Wizard 2480 automatic γ-counter (Perkin Elmer, Inc.). The measured activity concentrations were converted to percentage injected activity per kilogram (%ID/kg). Metabolite analysis of activity in plasma was performed by reversed-phase HPLC with in-line radiation (Posi-RAM model 4, LabLogic) detection using a Kinetex Biphenyl column (Phenomenex, 150 × 4.6 mm; 5 µm particle size) and a mobile phase gradient of 10–75% acetonitrile (0.1% TFA) in water (0.1% TFA) over 20 minutes. Intact ^18^F-PARPi elutes at 16 minutes and a number of metabolites elute from 7–8 minutes.

### Dosimetry

Absorbed radiation doses to normal tissues were estimated for the first 6 patients of the study, based on dynamic and static PET/CT images. Activity concentration-time curves were generated by analysis of VOIs generated for liver, kidney, spleen, cardiac blood pool, bone, lung, gallbladder and urinary bladder. Red marrow activity concentration was assumed equal to that of blood. WB activity-time curves, generated using the 4 points defined by the administered activity (time zero) and the total activities in the three whole-body PET scans, were used to calculate mono-exponential clearance half-times. The areas under activity concentration-time curves (AUC) were estimated by trapezoidal integration with a terminal contribution calculated by extrapolation from the last measured value using the shorter of apparent terminal clearance rate or physical decay. Whole-organ AUCs were obtained by multiplying the activity concentration AUC by organ mass. Baseline values of organ mass were taken from the Oak Ridge National Laboratory (ORNL) phantoms of OLINDA/EXM® 2.0 (Hermes Medical Solutions, Sweden) representing standard human. Organ masses were rescaled if body mass differed by more than 15% from the standard value (73.7 kg for males; 56.9 kg for females). Organ residence times were derived by dividing organ AUC values by administered activity. For urinary bladder contents, residence times were estimated by the OLINDA/EXM 2.0 voiding bladder model based on the fraction of activity clearing via urinary bladder, the mono-exponential whole-body biological half-time, and an assumed voiding interval of 1 hour. Residence times for the remainder of body were derived by subtracting all the individually estimated residence times from the whole-body residence time. Absorbed radiation doses to the whole body and various organs were calculated using OLINDA/EXM 2.0 with effective doses based on the tissue weighting factors of ICRP Report 103 (26).

### PET/CT imaging and analysis

All PET/CT images were obtained on a Discovery 710 PET/CT scanner (GE Healthcare), using low dose CT settings (10-80 mA, 120 kV) for CT images that were used for attenuation correction and anatomic correlation. All studies were reviewed using the Hybrid Viewer display and analysis application (Hermes Medical Solutions, Sweden). ^18^F-PARPi PET/CT and ^18^F-FDG PET/CT studies were interpreted by two nuclear medicine physicians with at least 10 years of PET/CT experience. Three-dimensional threshold-based volumetric regions of interest (VOI) were placed in reference regions (bilateral submandibular gland, parotid gland, blood pool of neck, contralateral posterior neck muscle, genioglossus, bone marrow, mediastinal blood pool, myocardium, normal liver, renal cortex, and spleen) and over all sites of abnormal uptake in lymph nodes, or soft-tissue lesions with reference to the PET/CT images. Abnormal ^18^F-PARPi and ^18^F-FDG uptake was defined as outside physiologic sites (such as palatine tonsils or skeletal muscle) and of intensity greater than regional background. Uptake of ^18^F-PARPi in the soft-tissue lesions and lymph nodes was assessed by measuring the maximum standardized uptake values (SUV_max_).

### Statistics

Uptake values are presented as mean ± standard-error, unless otherwise specified. Distribution of uptake with ^18^F-PARPi and ^18^F-FDG (on lesions where both were available) were compared using a Mann-Whitney-Wilcoxon test for paired data, while their variances were compared using a Levene’s test. R version 3.6.0 was used for analysis.

## Results

### Patient population

A total of 11 patients with cytologically or histologically confirmed squamous cell carcinoma of the oral cavity or oropharynx completed the study protocol. Table 1 lists the patient demographics and key diagnostic parameters, including stage and lymph node status, and Figure 1 shows a schematic overview of the study workflow. There were 10 males and one female, with a mean age of 64 years. At the time of imaging, 8 patients had newly diagnosed disease (all with the primary lesion in the oropharynx). The 3 patients that had recurrent disease at the time of imaging had previously been diagnosed with oral cavity squamous cell carcinoma. Nodal involvement (anatomically abnormal lymph nodes) was present in 9 out of the 11 patients, and disease stage ranged from I to IVb (8^th^ edition AJCC). 27% of patients (n = 3) were HPV-negative (all oral cavity squamous cell carcinoma) and 73% were HPV-positive (all oropharyngeal cases). Six patients were smokers (5 of them with more than a 10 pack/year smoking history and 1 of them with less).

**Table 1.** Summary of patient and tumor characteristics, including key diagnostic parameters

| Patient No. | Age | Sex | Primary | Recurrent | Stage | Nodal status | HPV status | >10 y smoking |
| --- | --- | --- | --- | --- | --- | --- | --- | --- |
| 1 | 52 | M | oropharynx | no | I | N1 | positive | never |
| 2 | 72 | F | oral | yes | I | N0 | negative | never |
| 3 | 72 | M | oropharynx | yes | I | N1 | positive | yes |
| 4 | 53 | M | oropharynx | no | II | N0 | positive | no |
| 5 | 86 | M | oral | yes | IVa | N0 | negative | yes |
| 6 | 59 | M | oropharynx* | no | I | N1 | positive | never |
| 7 | 73 | M | oropharynx | no | I | N1 | positive | yes |
| 8 | 67 | M | oropharynx | no | II | N1 | positive | yes |
| 9 | 64 | M | oropharynx | no | I | N1 | positive | yes |
| 10 | 49 | M | oral | yes | IVb | N3 | negative | yes |
| 11 | 56 | M | oropharynx | yes | II | N1 | positive | no |

\* Not otherwise specified.

**Figure 1.**
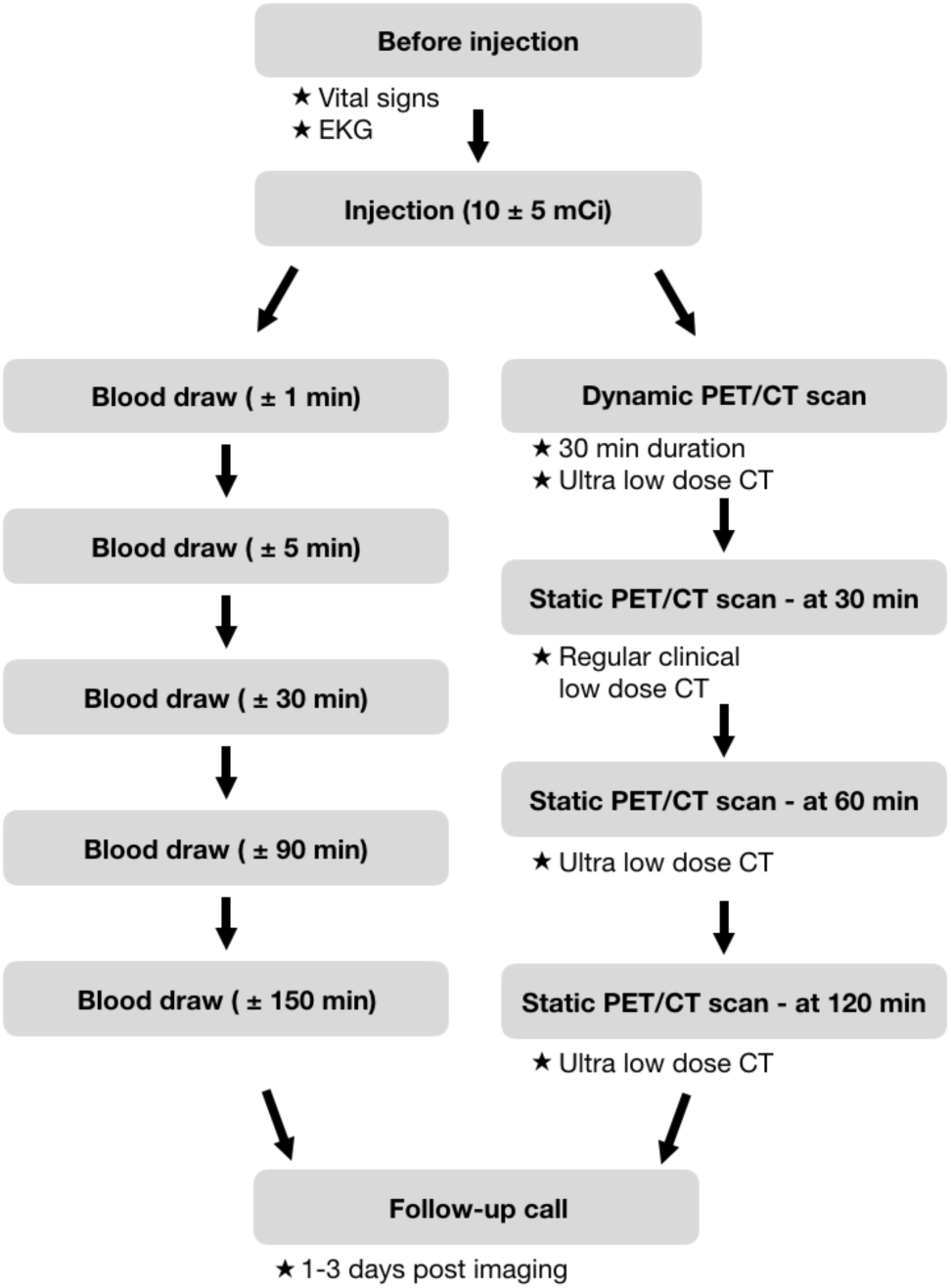
Schematic overview and flow chart of the ^18^F-PARPi Phase I clinical trial. Twelve patients were enrolled on this study protocol. Of these, eleven patients completed the study. One patient withdrew consent before administration of ^18^F-PARPi. No patients were excluded from the analysis.

### Radiotracer characteristics

The radiochemical synthesis of ^18^F-PARPi was performed using a synthetic route similar to what we have reported before (18-20,24). We first synthesized *para*-[^18^F]fluoro-benzoic acid as a radiolabeled synthon before conjugation with the 1(2H)phthalazinone targeting group. The molar activity was 115 ± 48 GBq/μmol. On average, patients were therefore injected with 290 pmol of ^18^F-PARPi, 6.7 orders of magnitude lower than the bi-daily administered dose of olaparib (2 × 300 mg) and therefore unlikely to elicit a pharmacodynamic response.

### Adverse events and metabolism

All patients tolerated the injection of ^18^F-PARPi well, and no adverse events were recorded related to the ^18^F-PARPi injection. One patient died within a 2-week window after completing the study, and one patient experienced grade 1 mucositis over the tumor site, which resolved the following day. The death was considered unrelated to the administered drug. The mucositis was considered possible related to the administered drug. For all patients, clinical chemistry and hematology were determined and an electrocardiogram was performed before and after the administration of ^18^F-PARPi (Supplementary Table S2). While some patients presented with abnormal findings before imaging, no clinically relevant changes were observed after radiotracer injection or at follow-up.

### Biodistribution and dosimetry

Maximum intensity projection PET images of a representative patient injected with ^18^F-PARPi (images obtained at 30 min, 60 min and 120 min p.i.) are shown in Figure 2A. At 120 min, uptake in the primary tumor had an SUV_max_ of 4.1, whereas lymph nodes had and SUV_max_ of 3.6 (level 1), 2.6 (level 2) and 2.4 (level 3). Across the entire patient population, the average primary tumor SUV_max_ was 2.8 ± 1.1 at 120 min. The primary routes of excretion were renal and hepatobiliary with most of the tracer excreted renally. Activity in the renal cortex diminished over time (SUV_max_ = 16 ± 8 at 30 min, 9 ± 4 at 60 min and 7 ± 5 at 120 min) with commensurate accumulation in the urinary bladder. The maximal observed activity in the urinary bladder corresponded to 20-38% of the total administered, typically at the 30-60 min scan times. Simultaneously a fraction of the tracer (on average 5% of the total administered) was excreted via the hepatobiliary route with relatively rapid passage through the liver (SUV_max_ = 7 ± 2 at 30 min, 6 ± 1 at 60 min, 4 ± 1 at 120 min), typically high accumulation in the gallbladder (SUV_max_ = 130 ± 92 at 120 min) and subsequent transit into the gut.

**Figure 2.**
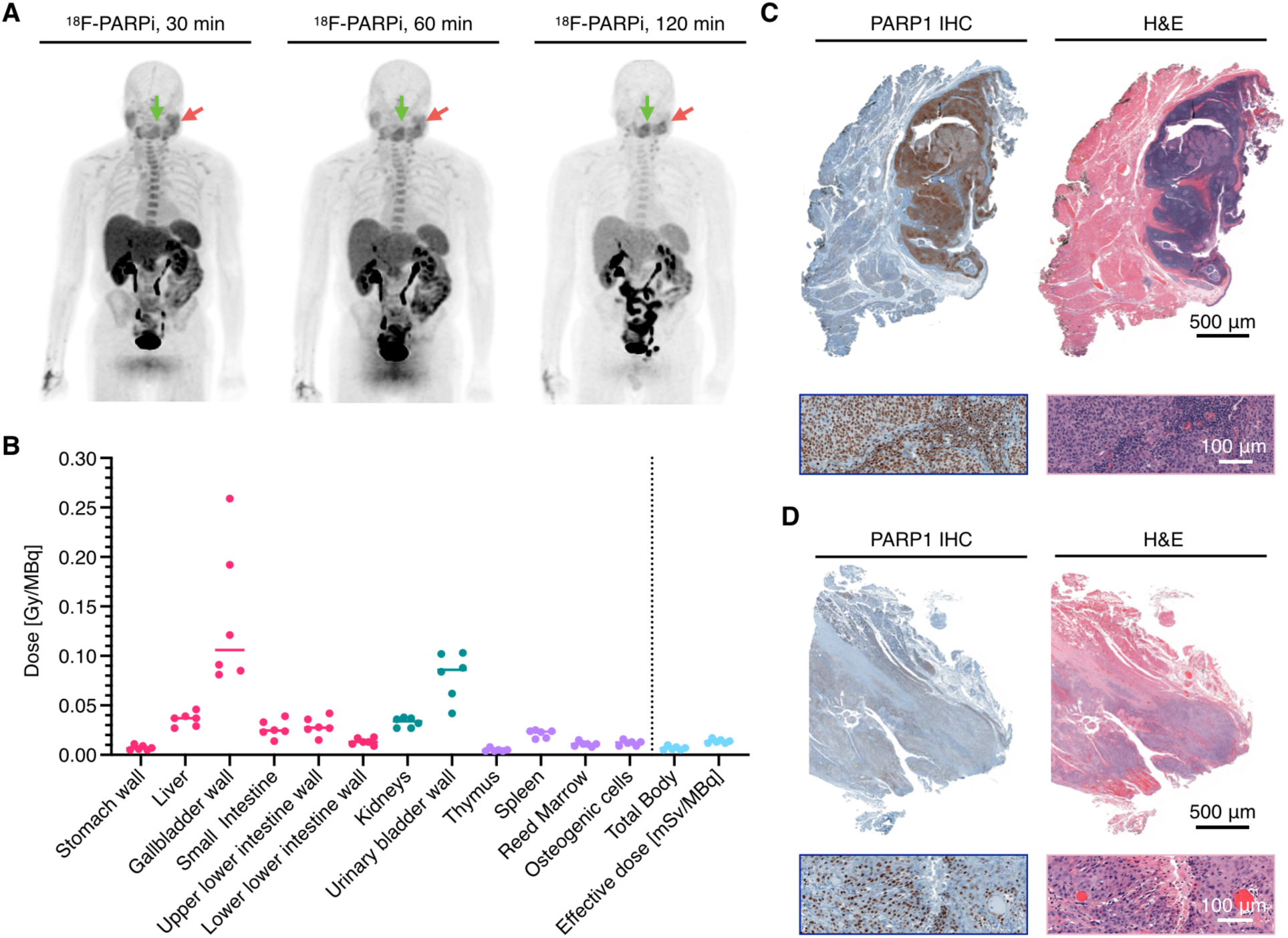
Biodistribution, histology and dosimetry of ^18^F-PARPi. (**A**) Maximum intensity projections at 30 min, 60 min and 120 min post ^18^F-PARPi injection in a 67 year old male with oropharyngeal squamous cell carcinoma. Green arrow: primary tumor; Red arrow: metastatic lymph node. (**B**) Dosimetry of select organs, whole body and effective dose in patients (n = 6) injected with ^18^F-PARP (see Supplementary Table S3 for the comprehensive dataset). (**C**) PARP1 IHC and corresponding H&E images of the primary lesion of a 49 year old male with carcinoma in the right palatine tonsil and corresponding H&E images. (**D**) PARP1 IHC and corresponding H&E images of the primary lesion of a 51 year old male with oral tongue carcinoma.

Absorbed radiation doses to normal tissues from ^18^F-PARPi were estimated based on the tracer biodistribution of the first six patients. Key dosimetry data are plotted in Figure 2B, and the entire dataset can be found in Supplementary Table S3. The effective dose of ^18^F-PARPi was 0.014 ± 0.002 mSv/MBq, calculated with ICRP103. The tissue compartments receiving the highest dose levels were the gallbladder wall (0.14 ± 0.07 mGy/MBq) and urinary bladder wall (0.08 ± 0.02 mGy/MBq). Radiation doses to bone marrow (0.011 ± 0.002 mGy/MBq), kidney (0.033 ± 0.005 mGy/MBq) and liver (0.036 ± 0.007 mGy/MBq) were considerably lower. In a typical diagnostic setting (280 MBq – 370 MBq ^18^F-PARPi injected intravenously), the effective radiation dose is projected to be 3.9 mSv – 5.2 mSv.

PARP1 was highly expressed in both oral and oropharyngeal squamous cell carcinoma in our patient population (Fig. 2C and D), corroborating earlier work (12,13).

### Metabolism

Research blood draws were obtained for 10 patients at five timepoints after tracer injection, activity counted and metabolites analyzed (Fig. 3A). Using a 2-phase decay curve, we determined the weighted blood half-life to be 4.2 min (Fig. 3B). Only small quantities of metabolites were detected at 1 min and 5 min (99.2 ± 1.5% and 89.9 ± 11.4% ^18^F-PARPi, respectively, Fig. 3C). At 30 min, and with decreasing blood pool concentration of the injected tracer, we detected a radiometabolite with a retention time of 7-8 min (50.9 ± 11.5%). For patients with blood samples at 90 min (n = 2) and 150 min (n = 1), increasing amounts of the metabolite were observed (58.8% and 72.1%, respectively). When screening possible non-radioactive, ^19^F-labeled degradation product analogues, we found that 4-fluorophenyl(4λ^2^-piperazin-1-yl)methanone had a retention time which was overlapping with the radiometabolite (t_R_ = 7.80 min). The small molecule therefore represented a likely component of the degradation pathway (Supplementary Fig. S2). This is consistent with the available literature on olaparib degradation (27).

**Figure 3.**
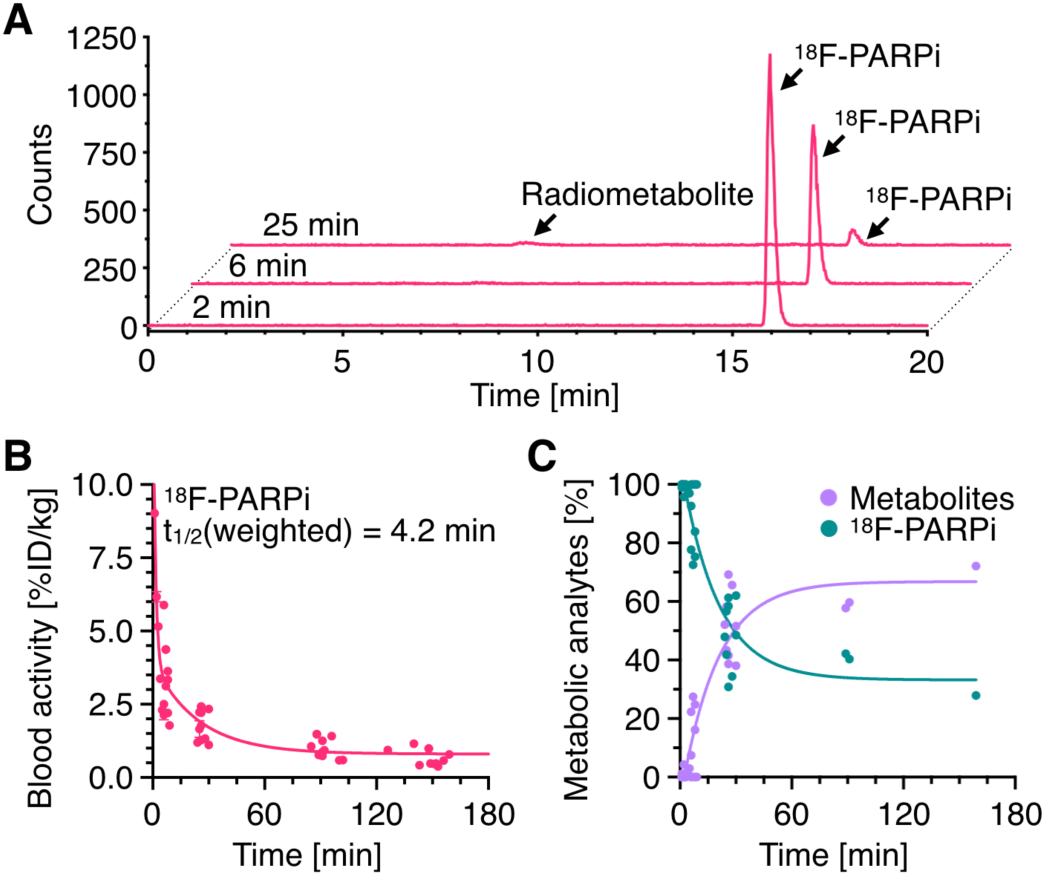
Metabolic stability and blood half-life ^18^F-PARPi in patients with oral and oropharyngeal cancer (n = 10). (**A**) HPLC chromatogram of ^18^F-PARPi in human blood serum of a patient at various time points post injection (2 min, 6 min and 25 min). (**B**) Weighted blood half-life of ^18^F-PARPi across all available patients (2-phase decay, no constraints, n = 10 patients). (**C**) Percentage of ^18^F-PARPi and metabolites at various time points post injection.

### Organ residence time

Investigating the specificity of ^18^F-PARPi, we looked at the residence times of the tracer in tumors, metastatic nodes and healthy tissues (Fig. 4). In spleen and liver, which both express large physiological amounts of PARP1 and PARP2 (28), initial uptake was high, followed by rapid clearance over the 2-hour imaging period (SUV_max_(spleen, 30 min) = 6.1 ± 1.3 and SUV_max_(spleen, 120 min) = 2.2 ± 0.6, representing a 64% drop). Similarly, the SUV_max_ in bone marrow was high in the 30 min PET/CT scan, but values declined by 53% between 30 mins and 120 mins. Comparably fast clearance was found for physiologic structures within the head and neck region. Uptake in the submandibular and parotid glands decreased by 57% and 56%, respectively (SUV_max_(submandibular, 30 min) = 3.6 ± 1.0 and SUV_max_(parotid, 30 min) = 3.1 ± 0.6; SUV_max_(submandibular, 120 min) = 1.6 ± 0.4 and SUV_max_(parotid, 120 min) = 1.4 ± 0.4). In contrast, tracer retention in tumor and metastatic nodes was significantly longer, with SUV_max_ values declining by just 13% for both primary and PET-avid lymph nodes (SUV_max_(tumor, 30 min) = 3.4 ± 0.8 and SUV_max_(tumor, 120 min) = 3.0 ± 1.1; SUV_max_(lymph node, 30 min) = 3.3 ± 1.3 and SUV_max_(lymph node, 120 min) = 2.9 ± 1.1).

**Figure 4.**
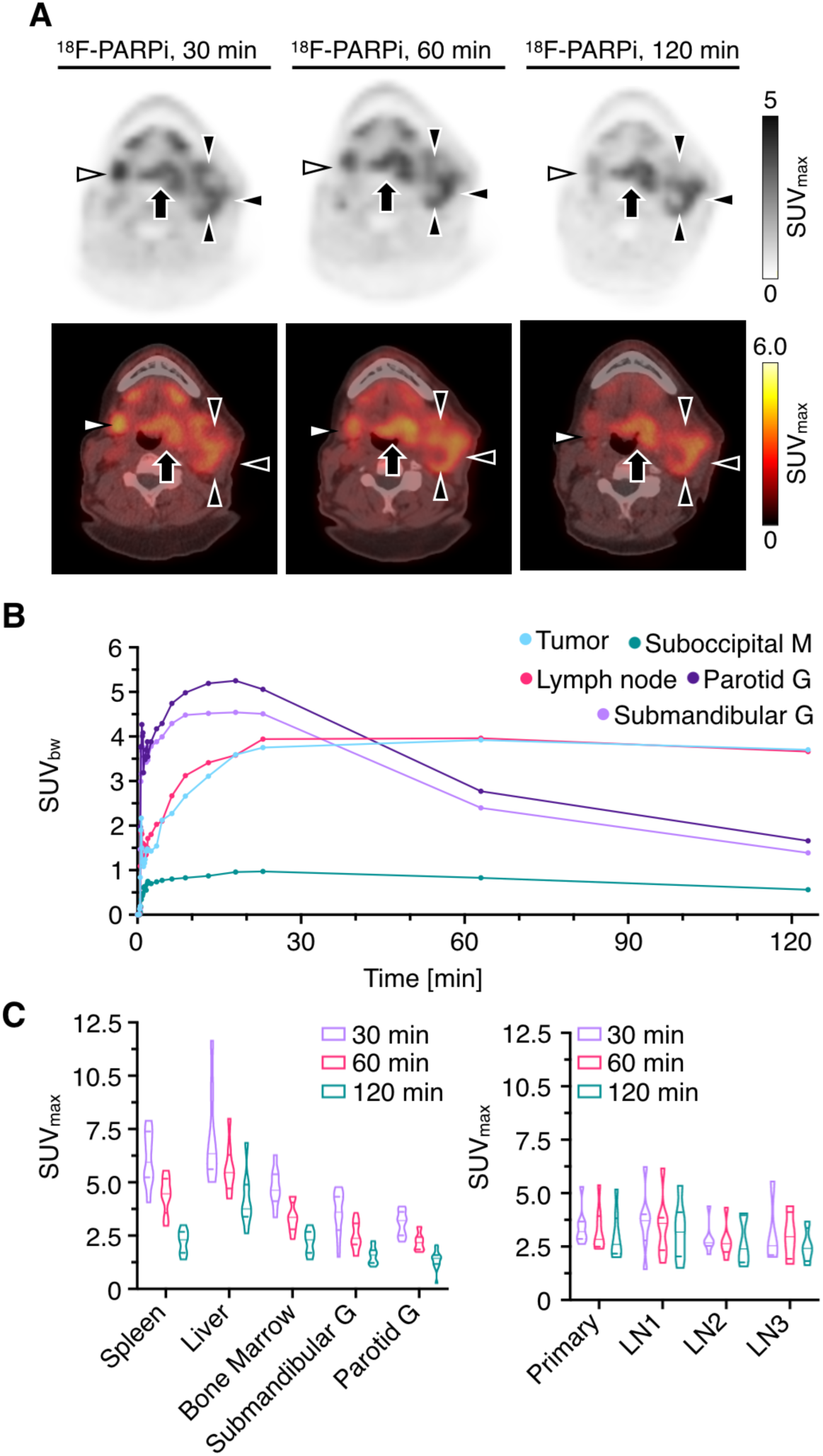
^18^F-PARPi pharmacokinetics in physiologic tissue, ^18^F-PARPi avid lymph nodes and primary tumor. (**A**) ^18^F-PARPi tracer accumulation in a 67 year-old-male with oropharyngeal squamous cell carcinoma is initially high in the parotid gland (white triangles) and subsequently decreasing. In comparison, the primary tumor in the base of the tongue (black arrow) and metastatic lymph node (black triangles) exhibit more sustained retention of the radiotracer. Top row: PET image only; Bottom row: PET/CT. (**B**) Time-activity curves of a 67-year old male patient, derived from the area with highest ^18^F-PARPi uptake (SUV_max_, primary tumor and metastatic lymph node) or from the whole structure (SUV_mean_, parotid gland, submandibular gland and suboccipital muscle). (**C**) SUV_max_ uptake values for normal structures (left), and primary tumor/metastatic lesions (right, each n = 6 patients with whole-body PET scans).

### ^18^F-FDG and ^18^F-PARPi in primary tumor and normal neck tissues

To determine if ^18^F-PARPi could be a relevant imaging tracer for the head and neck region, we compared its retention with that of standard of care ^18^F-FDG (Figure 5). Visually, ^18^F-PARPi and ^18^F-FDG appeared to have similar contrast ratios at 120 minutes post-injection (Fig. 5A and B). For both imaging agents, uptake was corroborated with tumor outlines defined by standard of care T1-weighted Gd-MRI imaging (Fig. 5C). Across the patient population, ^18^F-FDG had higher average tumor SUV_max_ values than ^18^F-PARPi, but SUV_max_ values for ^18^F-FDG decreased for level 2 and level 3 lymph nodes. A smaller decrease in SUV_max_ values was found for ^18^F-PARPi (66% and 15% for ^18^F-PARPi and ^18^F-FDG, respectively, when grouping primary/level 1 lymph nodes and level 2/level 3 lymph nodes; Fig. 5D). Interestingly, when comparing uptake ratios (SUV_max_(lesion)/SUV_max_(genioglossus)), we found similar median values for ^18^F-FDG and ^18^F-PARPi (median = 3.0 versus 3.3, p = 0.23), although the variance was less for ^18^F-PARPi than for ^18^F-FDG (1.3 versus 6.0, p = 0.001, Fig. 5E and F).

**Figure 5.**
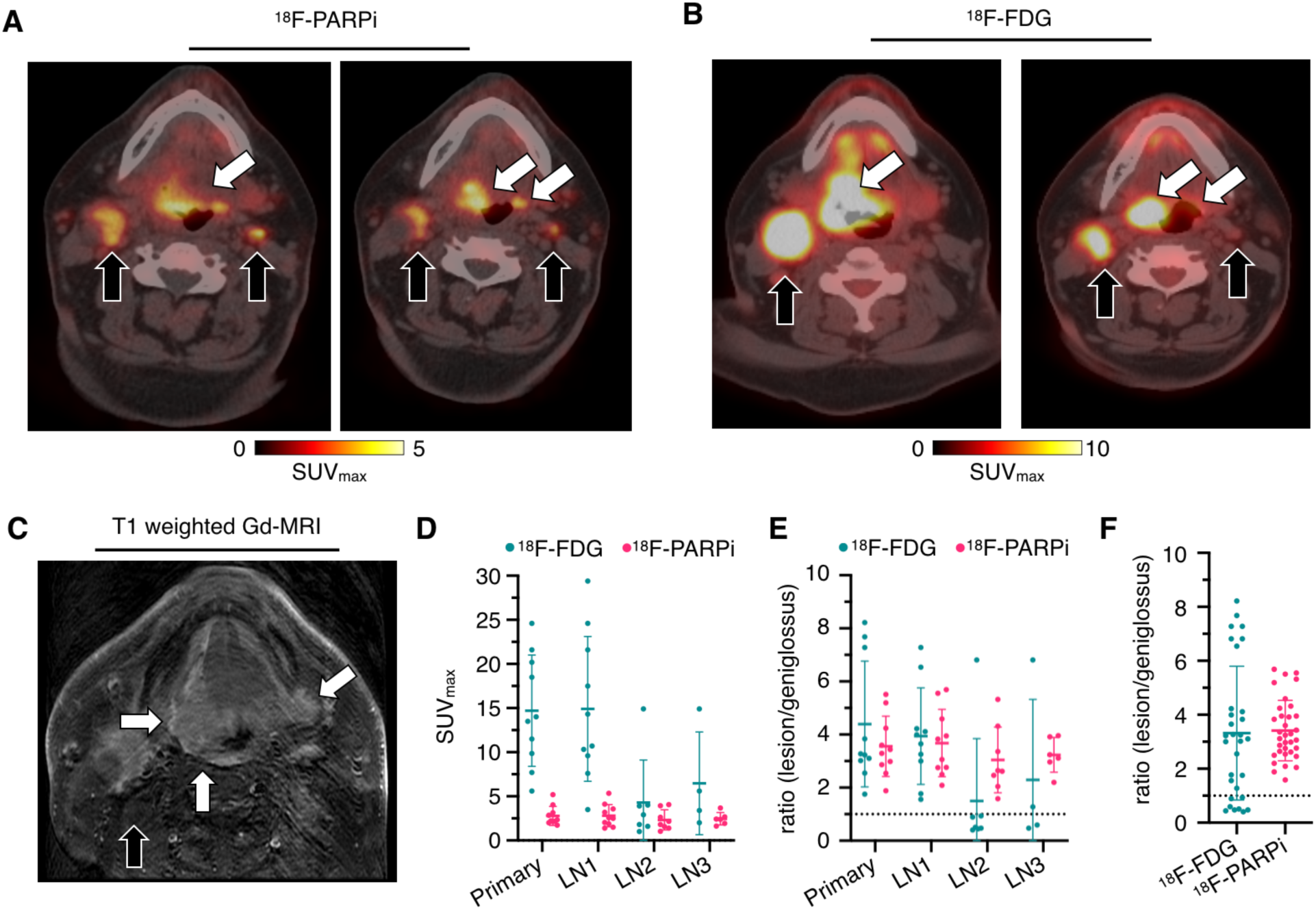
Comparison of standard of care ^18^F-FDG and ^18^F-PARPi in a 52 year-old-female with tongue cancer in base of the tongue. (**A**) ^18^F-PARPi PET showed an ^18^F-PARPi avid primary lesion in base of the tongue (white arrows) and metastatic lymph nodes (black arrows) at 120 min after the ^18^F-PARPi administration. (**B**) ^18^F-FDG PET also showed a ^18^F-FDG avid lesion in the base of the tongue (white arrows) and metastatic lymph nodes (black arrows). (**C**) Post contrast T1 weighted axial MRI imaging showed a that large expansile multicompartmental heterogeneously, ill-defined enhancing lesion centered in the base of tongue with effacement of the bilateral valleculae and extension along the median and lateral glossoepiglottic folds into the epiglottis. (**D**) Comparison of SUV_max_ values obtained for ^18^F-PARPi versus ^18^F-FDG in tumors and lymph nodes across the patient population. (**E**) SUV_max_-based uptake ratios for ^18^F-PARPi and ^18^F-FDG, separated by lesion location. (**F**) SUV_max_-based uptake ratios for ^18^F-PARPi and ^18^F-FDG without separation of primary lesion and lymph nodes. LN1 = ^18^F-FDG avid lymph node level 1; LN2 = ^18^F-FDG avid lymph node level 2; LN3 = ^18^F-FDG avid lymph node level 3.

^18^F-FDG and ^18^F-PARPi uptake matched with respect to the presence and location of the primary lesion. However, the two tracers had divergent uptake patterns in the lymphatic system (Fig. 6). For ^18^F-FDG, 34 lymph nodes were PET-avid. For ^18^F-PARPi, we observed 40 ^18^F-PARPi avid lymph nodes. These 40 lymph nodes included all of the lymph nodes that were detected using ^18^F-FDG. Due to protocol regulations, no biopsy material was available for the additional 6 lymph nodes, however, a subset of them resolved after chemoradiation (Fig. 6A and B).

**Figure 6.**
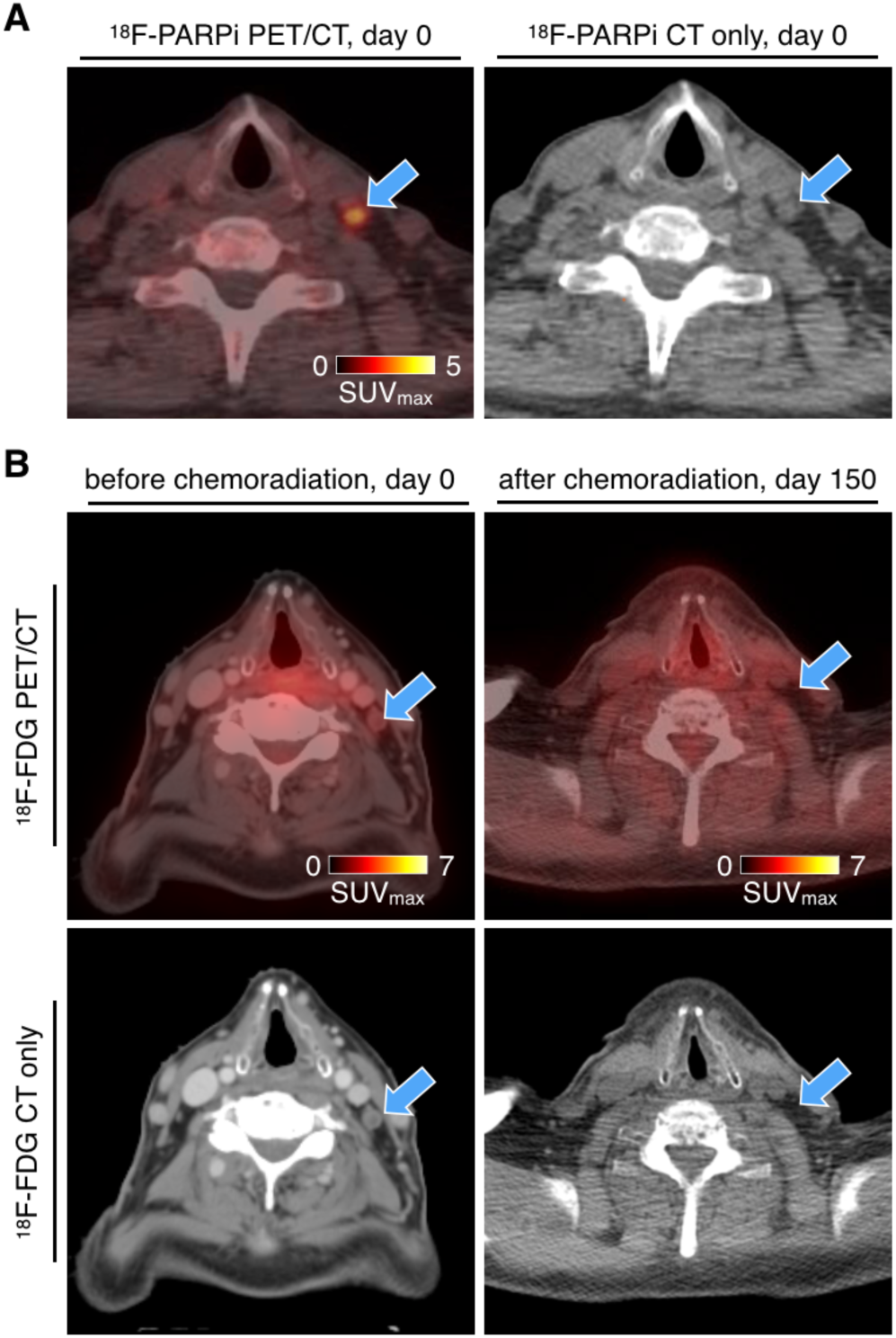
Post-treatment follow-up imaging in a 72-year-old male with tongue cancer in the left base of the tongue. (**A**) Pre-treatment ^18^F-PARPi PET/CT showed an ^18^F-PARPi-avid lesion in a neck lymph node (level 3, arrows) with an SUV_max_ value of 1.6 at 120 min after the ^18^F-PARPi administration. (**B**) Pre-treatment ^18^F-FDG PET/CT showed no ^18^F-FDG uptake in this lymph node (left column). Follow up imaging post chemoradiation showed that this lymph node had decreased in size. Blue arrows: ^18^F-PARPi avid and ^18^F-FDG negative lymph node.

## Discussion

Recently, PARP1/2-targeted agents have received considerable attention as imaging agents (16,29) based on the ubiquitous and near-universal expression of PARP in many types of cancers, with the promise to serve as an accurate sensor of malignancy where standard of care methods currently fail. This is of particular interest for head and neck imaging, where clinicians face multiple diagnostic obstacles, including the difficulty to correctly determine the extent of primary tumors (3,4) and the presence of metastatic disease (5,6). PARP1 is highly expressed in oral and oropharyngeal squamous cell carcinoma, and the percentage of PARP1-positive tissue in a biopsy specimen correlates with disease grade (13). Increased expression of PARP1 formed the foundation for this first-in-human study. Here we show that ^18^F-PARPi imaging is safe and feasible in a cohort of patients with biopsy-proven oral or oropharyngeal squamous cell carcinoma. PARP1/2-targeted ^18^F-PARPi uptake in primary lesions matched well with ^18^F-FDG uptake and MRI data, and high contrast images could be collected as soon as 120 min after injection. Our data further suggest that ^18^F-PARPi could have a higher detection rate of malignant lymph nodes and perhaps less nonspecific uptake in the head and neck region when compared to ^18^F-FDG.

Radiation doses associated with ^18^F-PARPi PET imaging were relatively low. The overall equivalent dose was 3.9 mSv – 5.2 mSv, lower than that reported for ^18^F-FDG (8.1 ± 1.2 mSv, (30)). This was in part due to the selective uptake of the tracer, paired with a short blood half-life and fast clearance. ^18^F-PARPi rapidly cleared from the circulatory system and accumulated in the urinary bladder and gallbladder following respective transit through the kidneys and liver. Kidney activity levels fell by 53% and liver by 41% between 30 mins and 120 mins of imaging. The gallbladder wall received the highest radiation dose, although with considerable variability (Fig. 2B) possibly due to the patients’ fasting status. Residence time in gallbladder, and hence radiation dose to gallbladder wall, could possibly be reduced by administration of CCK, a drug commonly used in clinical nuclear medicine practice (31). The urinary bladder wall received the second-highest effective dose, and could possibly be reduced, if necessary, by frequent bladder emptying.

PARP1/2 is located in the nuclei of cells. Consequently, the cell membrane and nuclear membrane permeability of ^18^F-PARPi has to be high, allowing both fast uptake of radiotracer and clearance of unbound material for image contrast generation. This was seen in the submandibular lymph nodes, where the SUV_max_ at 20-30 min was higher than in the primary tumor for 80% (n = 10 evaluable patients) of all patients (Fig. 4). Subsequently, the delivered activity in these healthy tissues cleared quickly, whereas the activity persisted longer in tumor and metastatic nodes (Fig. 4B). At 120 minutes post-injection the situation had inverted, and none of the submandibular lymph nodes showed activity higher than the primary tumor. Similar rapid clearance of radiotracer was noted in other healthy organs, including the spleen, genioglossus muscles, parotid glands and bone marrow. For future studies, imaging with PARP inhibitors after longer time intervals post-injection might yield further improved contrast ratios.

Our study has some limitations. Since this is a phase I study, we were unable to collect histological data from a statistically meaningful number of biospecimen for both primary and metastatic sites. Consequently, histologic diagnoses for cases where ^18^F-PARPi and ^18^F-FDG PET disagreed are not available, and while our data suggests that the tumor detection rate of ^18^F-PARPi is higher than the detection rate of ^18^F-FDG, it remains unclear if sensitivity and specificity are improved. Determining this – and consequently validating the clinical rationale for ^18^F-PARPi – will be the focus of a phase II study.

In conclusion, we performed the first-in-human translation of ^18^F-PARPi, a PARP1/2-targeted 1(2H)phthalazinone, and the first imaging of PARP1/2 in head and neck cancer. Administration of ^18^F-PARPi was safe, and aside from a grade 1 mucositis, which was possibly related, no adverse events were attributed to the tracer injection. ^18^F-PARPi is a promising new agent for the imaging of head and neck squamous cell carcinoma.

## Data Availability

Supplementary data is available online or from the corresponding authors.

## Author Contributions

H.S. and T.R. conceptualized and designed the study. H.S. and T.R. provided financial support. H.S. provided administrative support. S.R. and C.B. provided ^18^F-PARPi precursor. E.B., S.L. and J.S.L provided ^18^F-PARPi. A.M. provided biostatistical support. R.G. provided pathology support. R.N., M.P.D., I.G., S.P., N.Y.L and H.S. collected and assembled the data. H.S., P.D.D.S.F., R.N., M.G., J.O. and T.R. analyzed and interpreted the data. H.S. and T.R., with the help of all authors, wrote the manuscript. All authors approved the final manuscript.

## Acknowledgements

We acknowledge and thank Stephen Carlin and Kevin Staton for help with clinical radiochemistry; Aisha Shickler and Yorann Roux for blood and metabolite analysis; Ryan Min for patient coordination; Christopher Riedl, MD, PhD, for help with the clinical workflow; Susanne Kossatz, PhD and Wolfgang A. Weber, MD for helpful discussions and help with preparing the clinical trial.

## References

1. Zanoni DK, Montero PH, Migliacci JC, Shah JP, Wong RJ, Ganly I, et al. Survival outcomes after treatment of cancer of the oral cavity (1985-2015). Oral oncology 2019;90:115–21 doi 10.1016/j.oraloncology.2019.02.001.

2. Ho AS, Kim S, Tighiouart M, Gudino C, Mita A, Scher KS, et al. Metastatic Lymph Node Burden and Survival in Oral Cavity Cancer. Journal of clinical oncology : official journal of the American Society of Clinical Oncology 2017;35(31):3601–9 doi 10.1200/jco.2016.71.1176.

3. Gupta P, Migliacci JC, Montero PH, Zanoni DK, Shah JP, Patel SG, et al. Do we need a different staging system for tongue and gingivobuccal complex squamous cell cancers? Oral Oncol 2018;78:64–71 doi 10.1016/j.oraloncology.2018.01.013.

4. Zanoni DK, Migliacci JC, Xu B, Katabi N, Montero PH, Ganly I, et al. A Proposal to Redefine Close Surgical Margins in Squamous Cell Carcinoma of the Oral Tongue. JAMA Otolaryngol Head Neck Surg 2017;143(6):555–60 doi 10.1001/jamaoto.2016.4238.

5. Kim SJ, Pak K, Kim K. Diagnostic accuracy of F-18 FDG PET or PET/CT for detection of lymph node metastasis in clinically node negative head and neck cancer patients; A systematic review and meta-analysis. American journal of otolaryngology 2019;40(2):297–305 doi 10.1016/j.amjoto.2018.10.013.

6. Vali R, Bakkari A, Marie E, Kousha M, Charron M, Shammas A. FDG uptake in cervical lymph nodes in children without head and neck cancer. Pediatric radiology 2017;47(7):860–7 doi 10.1007/s00247-017-3835-8.

7. Rulach R, Zhou SY, Hendry F, Stobo D, James A, Dempsey MF, et al. 12 week PET-CT has low positive predictive value for nodal residual disease in human papillomavirus-positive oropharyngeal cancers. Oral Oncology 2019;97:76–81 doi 10.1016/j.oraloncology.2019.08.011.

8. Wang K, Wong TZ, Amdur RJ, Mendenhall WM, Sheets NC, Green R, et al. Pitfalls of post-treatment PET after de-intensified chemoradiotherapy for HPV-associated oropharynx cancer: Secondary analysis of a phase 2 trial. Oral Oncology 2018;78:108–13 doi 10.1016/j.oraloncology.2018.01.023.

9. D’Cruz AK, Vaish R, Kapre N, Dandekar M, Gupta S, Hawaldar R, et al. Elective versus Therapeutic Neck Dissection in Node-Negative Oral Cancer. N Engl J Med 2015;373(6):521–9 doi 10.1056/NEJMoa1506007.

10. Fasunla AJ, Greene BH, Timmesfeld N, Wiegand S, Werner JA, Sesterhenn AM. A meta-analysis of the randomized controlled trials on elective neck dissection versus therapeutic neck dissection in oral cavity cancers with clinically node-negative neck. Oral Oncol 2011;47(5):320–4 doi 10.1016/j.oraloncology.2011.03.009.

11. Mehanna H, Wong WL, McConkey CC, Rahman JK, Robinson M, Hartley AG, et al. PET-CT Surveillance versus Neck Dissection in Advanced Head and Neck Cancer. N Engl J Med 2016;374(15):1444–54 doi 10.1056/NEJMoa1514493.

12. Kossatz S, Brand C, Gutiontov S, Liu JT, Lee NY, Gonen M, et al. Detection and delineation of oral cancer with a PARP1 targeted optical imaging agent. Sci Rep 2016;6:21371 doi 10.1038/srep21371.

13. Kossatz S, Pirovano G, Franca PDDS, Strome AL, Sunny SP, Zanoni DK, et al. PARP1 as a biomarker for early detection and intraoperative tumor delineation in epithelial cancers – first-in-human results. bioRxiv 2019:663385.

14. Scott CL, Swisher EM, Kaufmann SH. Poly (ADP-ribose) polymerase inhibitors: recent advances and future development. J Clin Oncol 2015;33(12):1397–406 doi 10.1200/JCO.2014.58.8848.

15. Pommier Y, O’Connor MJ, de Bono J. Laying a trap to kill cancer cells: PARP inhibitors and their mechanisms of action. Sci Transl Med 2016;8(362):362ps17 doi 10.1126/scitranslmed.aaf9246.

16. Carney B, Kossatz S, Reiner T. Molecular Imaging of PARP. J Nucl Med 2017;58(7):1025–30 doi 10.2967/jnumed.117.189936.

17. Wilson TC, Xavier MA, Knight J, Verhoog S, Torres JB, Mosley M, et al. PET Imaging of PARP Expression Using (18)F-Olaparib. J Nucl Med 2019;60(4):504–10 doi 10.2967/jnumed.118.213223.

18. Carney B, Kossatz S, Lok BH, Schneeberger V, Gangangari KK, Pillarsetty NVK, et al. Target engagement imaging of PARP inhibitors in small-cell lung cancer. Nature communications 2018;9(1):176 doi 10.1038/s41467-017-02096-w.

19. Tang J, Salloum D, Carney B, Brand C, Kossatz S, Sadique A, et al. Targeted PET imaging strategy to differentiate malignant from inflamed lymph nodes in diffuse large B-cell lymphoma. Proceedings of the National Academy of Sciences of the United States of America 2017;114(36):E7441–e9 doi 10.1073/pnas.1705013114.

20. Carney B, Carlucci G, Salinas B, Di Gialleonardo V, Kossatz S, Vansteene A, et al. Non-invasive PET Imaging of PARP1 Expression in Glioblastoma Models. Mol Imaging Biol 2016;18(3):386–92 doi 10.1007/s11307-015-0904-y.

21. Michel LS, Dyroff S, Brooks FJ, Spayd KJ, Lim S, Engle JT, et al. PET of Poly (ADP-Ribose) Polymerase Activity in Cancer: Preclinical Assessment and First In-Human Studies. Radiology 2017;282(2):453–63 doi 10.1148/radiol.2016161929.

22. Makvandi M, Pantel A, Schwartz L, Schubert E, Xu K, Hsieh CJ, et al. A PET imaging agent for evaluating PARP-1 expression in ovarian cancer. J Clin Invest 2018;128(5):2116–26 doi 10.1172/JCI97992.

23. Michel LS, Dyroff S, Brooks FJ, Spayd KJ, Lim S, Engle JT, et al. PET of Poly (ADP-Ribose) Polymerase Activity in Cancer: Preclinical Assessment and First In-Human Studies. Radiology 2019;291(1):271 doi 10.1148/radiol.2019194006.

24. Kossatz S, Carney B, Schweitzer M, Carlucci G, Miloushev VZ, Maachani UB, et al. Biomarker-Based PET Imaging of Diffuse Intrinsic Pontine Glioma in Mouse Models. Cancer research 2017;77(8):2112–23 doi 10.1158/0008-5472.Can-16-2850.

25. Dunphy MPS, Harding JJ, Venneti S, Zhang H, Burnazi EM, Bromberg J, et al. In Vivo PET Assay of Tumor Glutamine Flux and Metabolism: In-Human Trial of (18)F-(2S,4R)- 4-Fluoroglutamine. Radiology 2018;287(2):667–75 doi 10.1148/radiol.2017162610.

26. The 2007 Recommendations of the International Commission on Radiological Protection. ICRP publication 103. Ann ICRP 2007;37(2-4):1–332 doi 10.1016/j.icrp.2007.10.003.

27. Thummar M, Kuswah BS, Samanthula G, Bulbake U, Gour J, Khan W. Validated stability indicating assay method of olaparib: LC-ESI-Q-TOF-MS/MS and NMR studies for characterization of its new hydrolytic and oxidative forced degradation products. J Pharm Biomed Anal 2018;160:89–98 doi 10.1016/j.jpba.2018.07.017.

28. Uhlen M, Fagerberg L, Hallstrom BM, Lindskog C, Oksvold P, Mardinoglu A, et al. Proteomics. Tissue-based map of the human proteome. Science 2015;347(6220):1260419 doi 10.1126/science.1260419.

29. Knight JC, Koustoulidou S, Cornelissen B. Imaging the DNA damage response with PET and SPECT. Eur J Nucl Med Mol Imaging 2017;44(6):1065–78 doi 10.1007/s00259-016-3604-1.

30. Quinn B, Dauer Z, Pandit-Taskar N, Schoder H, Dauer LT. Radiation dosimetry of 18F- FDG PET/CT: incorporating exam-specific parameters in dose estimates. BMC Med Imaging 2016;16(1):41 doi 10.1186/s12880-016-0143-y.

31. Krishnamurthy GT, Krishnamurthy S, Brown PH. Constancy and variability of gallbladder ejection fraction: impact on diagnosis and therapy. J Nucl Med 2004;45(11):1872–7.

